# Unveiling the Hidden Syndrome: The Enigma of Anti-Transcobalamin Receptor Autoantibodies

**DOI:** 10.1101/2024.11.19.24317544

**Authors:** Kazuki M. Matsuda, Hirohito Kotani, Shinichi Sato, Ayumi Yoshizaki

## Abstract

The transcobalamin receptor (CD320) functions as a critical mediator for vitamin B12 uptake in cells, with emerging evidence linking autoantibodies against CD320 to various autoimmune conditions. Pluvinage *et al.*’s recent study identified anti-CD320 autoantibodies as a cause of autoimmune vitamin B12 central deficiency, specifically affecting the central nervous system while sparing peripheral nerves. Their findings align with our previous work showing anti-CD320’s role in cutaneous arteritis. Both studies identified overlapping CD320 epitopes targeted by autoantibodies and demonstrated the therapeutic efficacy of high-dose vitamin B12 supplementation in mitigating symptoms by reducing CD320 expression on the surface of vascular endothelial cells. Expanding on these findings, we observed anti-CD320 autoantibodies in systemic sclerosis, systemic lupus erythematosus, and other inflammatory disorders, suggesting a broader clinical relevance. The work by Pluvinage *et al.* and our group supports the concept of an "anti-CD320-associated syndrome," with high-dose B12 supplementation as a promising treatment strategy. Further research is needed to fully elucidate the tissue-specific mechanisms and pathophysiology underlying these autoimmune conditions.

## Introduction

Cutaneous arteritis (CA), or cutaneous polyarteritis nodosa (PN), is a single-organ vasculitis marked by necrotizing inflammation in small to medium-sized arteries, similar to PN.(1) While PN impacts multiple organs, CA is confined to the skin and nearby joints, muscles, and peripheral nerves.(2) Diagnosis typically relies on skin biopsy, a procedure that can be burdensome for both patients and clinicians. Treatment often involves systemic corticosteroids and immunosuppressants,(3) though responses vary among patients.(4) There is a need for easily measurable biomarkers to aid in diagnosis and prognosis, along with a deeper understanding of the disease’s pathophysiology to develop targeted therapies.

In our previous work, we reported the presence of autoantibodies targeting the extracellular domain of CD320 in 24% of patients with cutaneous arteritis (CA).(5) Transcobalamin receptor (CD320), a single-pass transmembrane protein, functions as the cell uptake receptor for vitamin B12 (VB12).(6) Patients with positive anti-CD320 antibodies (Abs) were spared from peripheral neuropathy compared to those with negative anti-CD320 Abs. Immunohistochemical analysis revealed CD320 expression in the endothelium of arterioles in CA-affected skin, suggesting a direct role for anti-CD320 in CA pathogenesis. Notably, we demonstrated that anti-CD320 can induce an autocrine loop of interleukin-6 via internalization into human dermal vascular endothelial cells through endocytosis *in vitro*, leading to periarterial inflammation in murine skin i*n vivo*. Furthermore, we demonstrated that methylcobalamin (MetCbl), one of active form of vitamin B12 (VB12) which has been widely used in clinics for the treatment of peripheral neuropathy and megaloblastic anemia, ameliorates anti-CD320 Ab-induced inflammation through internalization of CD320 on the cell surface of endothelial cells.

We were profoundly intrigued by the recent publication by Pluvinage *et al.* detailing the role of autoantibodies against CD320 in the etiology of autoimmune vitamin B12 central deficiency (ABCD).(7) Using programmable phage display, Pluvinage *et al.* identified anti-CD320 antibodies in some patients with central nervous system (CNS) deficits and neuropsychiatric lupus. These antibodies impaired the cellular uptake of VB12 *in vitro* by depleting CD320 from the cell surface. Immunosuppressive therapy and high-dose systemic VB12 supplementation were linked to increased VB12 levels in the cerebrospinal fluid and clinical improvement. Interestingly, ABCD cases with anti-CD320 Abs showed no hematologic signs of VB12 deficiency. A genome wide CRISPR screen revealed that the low-density lipoprotein receptor (LDLR) provides an alternative VB12 uptake pathway in hematopoietic cells. In addition, recent case reports have demonstrated anti-CD320 Ab positivity not only in CA but also in other conditions, such as systemic sclerosis (SSc).(8,9) This works have collectively expanded the potential scope of anti-CD320 Ab-associated pathology in humans.

In this study, we present our work on epitope mapping of anti-CD320 Abs and the hematological features in CA, discussing the alignment between findings in CA and ABCD. We also utilized publicly available single-cell RNA sequencing data to examine *CD320* expression in human endothelial cells across various tissues, exploring the tissue-specific nature of anti-CD320 Ab-associated pathology. Additionally, we highlight our latest research on proteome-wide autoantibody screening (PWAbS) across multiple human disorders, mapping the landscape of autoimmunity targeting CD320. Our aim is to compare the findings of Pluvinage *et al*. with our own, providing a comprehensive overview of the "anti-CD320-associated syndrome" in humans. We seek to highlight critical areas for future research and deepen the understanding of this condition.

## Materials and Methods

### Human subjects

We enrolled CA patients visited our clinic from April 2020 to November 2022, whose sera were available from our sample stock. All the patients fulfilled the diagnostic criteria suggested by Nakamura T et al. in 2009.(2) We also recruited patients with systemic sclerosis (SSc) from autoantigenome database named “autoantibody comprehensive database (UT-ABCD)”.(10) Clinical data were collected by retrospective review of electric medical records. The demographics of the participants have been described previously.(5) We gathered laboratory findings from the closest time point from the date of serum collection. This study has been approved by The University of Tokyo Ethical Committee (Approval number 2023051G). Written <u>informed consent</u> has been obtained from all the participants.

### Epitope mapping

We conducted epitope mapping for anti-CD320 Abs in CA cases utilizing wet protein arrays (WPAs) as previously described.(5,11,12) In brief, we synthesized truncated forms of CD320 protein from our entry clone (NCBI Reference Sequence: NM_016579.2) with GST and FLAG tags and linkers added on their N-terminus, utilizing a wheat germ cell-free system.(13) The synthesized proteins were attached to 96-well plates coated with glutathione by its affinity with GST tags. The antigen-coated plates were reacted with Alexa Fluor 647-conjugated anti-FLAG Ab (MBL, Tokyo, Japan) diluted by 1:3000 for 1 hour at room temperature. After washing the plate, fluorescence was measured by a fluorescence imager (Typhoon FLA 9500). Next, the plates were washed again and treated with human serum diluted by 1:150 for 1 hour at room temperature, and subsequently with Alexa Fluor 647-conjugated goat anti-human IgG (H+L) Ab (Thermo Fisher Scientific). After washing the plates, fluorescence was measured by a fluorescence imager (Typhoon FLA 9500). Fluorescence measurements obtained from each well were corrected by the following formula.

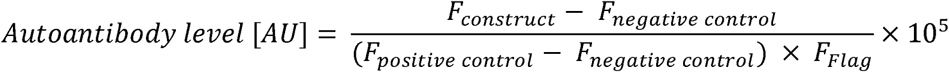

*AU*: arbitrary unit
*F _construct_*: fluorescent intensity of a spot coated by each construct
*F _negative control_*: fluorescent intensity of a null spot as a negative control
*F _positive control_*: fluorescent intensity of a spot coated by IgG as a positive control

### Database search

Expression of *CD320* in endothelial cells across multiple human tissues, as measured by single-cell RNA-sequencing, was referenced from the Tabula Sapiens project.(14) Serum levels of anti-CD320 Abs in various conditions including COVID-19, atopic dermatitis (AD), anti-neutrophil cytoplasmic antibody-associated vasculitis (AAV), systemic lupus erythematosus (SLE), SSc, and healthy controls (HCs) were cited from UT-ABCD.(10)

### Data visualization

The heatmap was visualized using GraphPad Prism. Box plots were made by R as follows: the middle line corresponds to the median; the lower and upper hinges correspond to the first and third quartiles; the upper whisker extends from the hinge to the largest value no further than 1.5 times the interquartile range (IQR) from the hinge; and the lower whisker extends from the hinge to the smallest value at most 1.5 times the IQR of the hinge.

### Statistical analysis

Statistical analyses were performed using R. P values were calculated by Mann Whitney U test.

## Results

### Epitope mapping

We have previously identified the major epitope of anti-CD320 Abs in CA as Thr^169^-Tyr^229^ employing WPAs.(5) Herein we conducted further mapping by displaying truncated forms of CD320 (**Figure 1A**) on the same WPA system, narrowing the epitope into Ser^189^-Thr^198^ (**Figure 1B**). We also performed epitope mapping on anti-CD320-positive sera from patients with SSc, revealing identical epitopes to those found in CA. This key epitope closely matched the one identified by Pluvinage *et al*., using a combination of phage display, sequential alanine mutagenesis, and immunoprecipitation, spanning Pro^183^-Thr^197^.(7)

**Figure 1.**
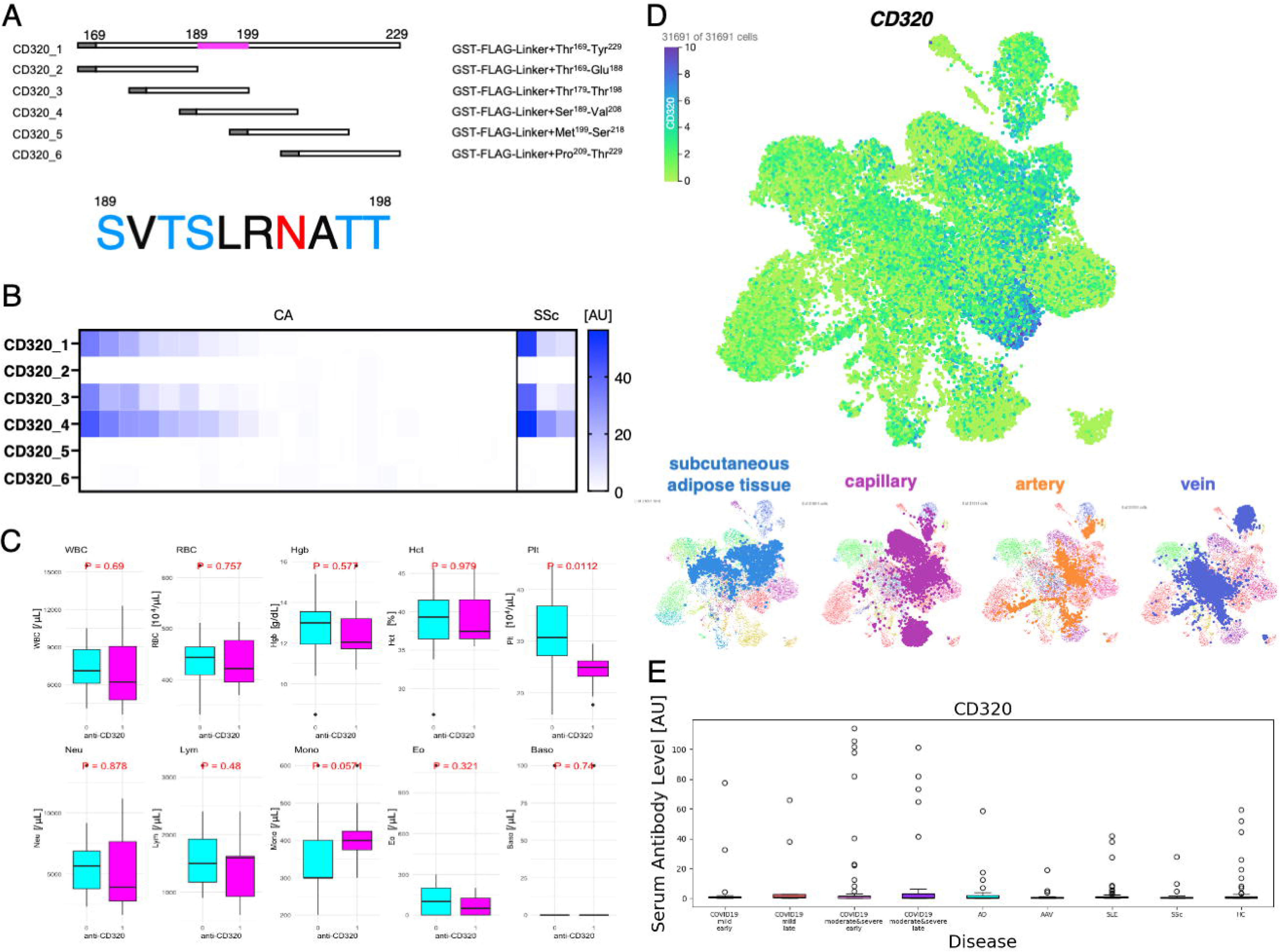
(A) Truncated forms of CD320 prepared for epitope mapping in detail. The amino acid sequence of identified major epitopes (magenta) is shown below. The epitope was rich in potential O-linked (aqua) or N- linked (red) glycosylation sites (B) Heat map illustrating signal strength from each spot upon wet protein arrays displaying truncated forms of CD320 treated with serum samples and fluorescence-conjugated secondary antibody. Each column represents a different human subject. AU: arbitrary unit. (C) Hematological features of CA patients by serum anti-CD320 seropositivity. WBC: white blood cell, RBC: red blood cell, Hgb: hemoglobin, Hct: hematocrit, Plt: platelet, Neu: neutrophil, Lym: lymphocyte, Mono: monocyte, Eo: eosinophil, Baso: basophil. P values are calculated by Mann-Whitney’s U test. (D) Expression levels of *CD320* at single-cell resolution in human endothelial cells from the Tabula Sapiens. Cell type and tissue annotations are also shown. (E) Serum levels of anti-CD320 Abs in various conditions as reported in UT-ABCD.

### Hematological features of CA

Our clinical data on CA patients showed no association between anti-CD320 positivity and hematological abnormalities, excluding lower platelet counts within standard range (31.3 × 10^4^±7.4 × 10^4^ vs 24.2 × 10^4^±3.9 × 10^4^, P = 0.01; **Figure 1C**).

### Single cell transcriptome analysis

While single-cell RNA sequencing analysis by Pluvinage *et al.* indicated elevated *CD320* expression in CNS endothelium, we have confirmed *CD320* expression in the endothelium of subcutaneous adipose tissue, the primary locus of CA, utilizing the Tabula Sapiens (**Figure 1D**).(14) Expression levels of *CD320* seemed to be similar between arteries and veins, and relatively low in capillaries.

### Anti-CD320 seropositivity in various conditions

We referred to our latest work involving PWAbS targeting a total of 284 human subjects with various inflammatory disorders, which unveiled the presence of anti-CD320 Abs in a wide range of conditions including SLE, SSc, and COVID-19, as well as in HCs (**Figure 1E**).(10)

## Discussion

In this study, we identified that the major epitope of anti-CD320 Abs in CA and SSc lies within Ser^189^-Thr^198^ (**Figure 1A and 1B**). No significant association was found between anti-CD320 seropositivity and hematological abnormalities in CA, except for low platelet counts within standard range (**Figure 1C**). Single-cell RNA sequencing data revealed that *CD320* is expressed in subcutaneous adipose tissue, arteries, and veins (**Figure 1D**). Additionally, data from UT-ABCD showed that anti-CD320 seropositivity is widely observed in various human conditions beyond CA, including COVID-19, SSc, SLE, and even in healthy individuals (**Figure 1E**).

Our findings align with those of Pluvinage *et al.* in several key aspects. First, the CD320 epitope targeted by autoantibodies in both studies was nearly identical (**Figure 1A and 1B**). The consistency of these major epitopes across different patient populations and detection methods reinforces the robustness of the identified epitope and its significance in human pathology. Second, in both CA and ABCD, the peripheral nervous system was unaffected in cases positive for anti-CD320 Abs. Pluvinage *et al*. proposed a model suggesting that anti-CD320 Abs specifically target the CNS because the CNS depends on CD320-mediated VB12 uptake beyond the blood-brain barrier, while peripheral nerves receive VB12 through passive transport.(7) Third, MetCbl showed a protective effect against anti-CD320 antibody-associated pathology in both studies by inducing internalization of CD320, thereby preventing it from being targeted by autoantibodies on the cell surface.(5,7) As a water-soluble vitamin, MetCbl rarely causes hypervitaminosis, and its safety in high doses has been confirmed in clinical trials for amyotrophic lateral sclerosis (ALS).(15) It has recently been approved in Japan as a treatment for early-stage ALS. The effectiveness of VB12 administration in both studies supports the pathogenic role of anti-CD320 Abs by binding to CD320 and suggests that VB12 supplementation could serve as a highly safe, targeted therapeutic strategy. Further validation through clinical interventional studies is needed as previously mentioned.(5)

There were also notable differences between the findings of Pluvinage *et al*. and our own. While Pluvinage *et al*. reported no hematological abnormalities in ABCD cases, in our CA cohort, anti-CD320 seropositivity was associated with low platelet counts, though still within the standard range (**Figure 1C**). It is important to note that isolated thrombocytopenia has been recognized as an uncommon clinical presentation of VB12 deficiency.(16) Additionally, Pluvinage *et al*. focused exclusively on a granulocyte-lineage leukemia cell line in their study,(7) suggesting that further investigation is required in other hematopoietic cell lineages.

Our analysis of the single-cell RNA sequencing data revealed that CD320 expression on endothelial cells is not limited to the CNS but is also found in subcutaneous adipose tissue, arteries, and veins (**Figure 1D**). This suggests that anti-CD320 Abs could potentially trigger both CA and ABCD even in the same individual. In fact, we recently reported a case with anti-CD320 seropositivity who experienced CA along with cranial nerve impairments of unknown etiology.(9) Moreover, some cases with CNS deficits presented by Pluvinage *et al*. showed focal neurological symptoms and brain imaging findings,(7) which are uncommon in typical VB12 deficiency but are often seen in CNS vasculitis.(17,18) These findings indicate that the coexistence of B12 deficiency and vasculitis, driven by anti-CD320 Abs, cannot be excluded, even in the cases described by Pluvinage *et al.*(7)

It is also important to highlight that both studies found some healthy individuals to be seropositive for anti-CD320 Abs (**Figure 1E**). The clinical significance of anti-CD320 Abs in healthy individuals, or in conditions other than CA and ABCD, remains unclear. To determine whether these autoantibodies play a direct role in disease development, exacerbate existing pathology, or are incidental, future studies should focus on prospective recruitment and longitudinal observation of seropositive patients. These findings suggest that the clinical relevance of anti-CD320 Abs may extend to a broader spectrum of conditions.

The tissue selectivity of CA and ABCD also remains a mystery, as it cannot be fully explained by the expression pattern of CD320 across different tissues. One hypothesis is the "two-hit theory," which posits that both the presence of autoantibodies and antigen exposure are necessary for disease manifestation. The major CD320 epitope we identified is rich in serine, threonine, and asparagine residues (**Figure 1A**), which are highly glycosylated in the human body.(19) Notably, the antigen expression systems used by both Pluvinage *et al*. and our team involved non-human cells,(5,7) meaning that proper glycosylation of the protein was not replicated. There is evidence that autoantibody seropositivity can depend on post-translational modifications like glycosylation.(20) Therefore, further investigation into how post-translational modifications, particularly glycosylation, affect the pathogenesis of anti-CD320 autoantibody-associated conditions is warranted.

Collectively, the research by Pluvinage *et al.* has made significant strides in understanding the role of anti-CD320 autoantibodies in VB12 deficiency, particularly within the CNS.(7) Their findings, in line with our own research,(5) highlight the potential of these autoantibodies in contributing to a spectrum of autoimmune conditions. Both studies suggest "anti-CD320-associated syndrome" as a novel disease concept. This idea provides new insights into the pathogenesis of autoimmune disorders in humans and highlights the potential of high-dose B12 supplementation as a novel, pathophysiology-oriented treatment with high safety and efficacy. The selectivity of the targeted tissues and the molecular mechanisms linking the autoantibodies to pathogenesis should be further investigated in depth.

## Data Availability

All data produced in the present study are available upon reasonable request to the authors.

## Acknowledgements

We thank Ms. Maiko Enomoto and her colleagues for their secretarial work. We appreciate K. Yamaguchi, T. Okumura, C. Ono, and N. Goshima from ProteoBridge Corporation for preparing the WPAs. We acknowledge R. Uchino, Y. Murakami, and H. Matsunaka from TOKIWA Pharmaceuticals Co. Ltd. for providing technical assistance with autoantibody measurement. We also thank E. Fukuda and A. Kuno from National Institute of Advanced Industrial Science and Technology for their valuable advice on protein glycosylation.

## Author Contributions

KM Matsuda primarily engaged in autoantibody measurement, collecting serum samples and clinical data, data analysis, visualization, and writing the first draft of the manuscript. H Kotani oversaw epitope mapping. S Sato conceptualized and supervised the study. A Yoshizaki conceptualized and supervised this study and was involved in revising the manuscript.

